# Detection of Subclinical Pulmonary Edema After Acute ST-segment Elevation Myocardial Infarction Without Heart Failure by Chest Computed Tomography

**DOI:** 10.1101/2023.03.04.23286664

**Authors:** Minghui Hua, Rong Liang, Yufan Gao, Keyi Cui, Yanzhen Liu, Shuo Liang, Ximing Li, Hong Zhang

**Affiliations:** Department of Radiology, Chest Hospital, Tianjin University, Tianjin, China; Department of Breast Imaging, Tianjin Medical University Cancer Institute and Hospital, National Clinical Research Center for Cancer, Tianjin, China; Academy of Medical Engineering and Translational Medicine, Tianjin University, Tianjin, China; Department of Cardiology, Chest Hospital, Tianjin University, Tianjin, China

**Keywords:** pulmonary edema, ST-segment elevation myocardial infarction, computed tomography, XGBoost, cardiac enzyme, prognosis

## Abstract

**BACKGROUND:** Pulmonary edema is a severe complication in patients with acute myocardial infarction which indicates the development of heart failure (HF) and poor prognosis. However, subclinical pulmonary edema after acute ST-segment elevation myocardial infarction (STEMI) without HF has not received enough attention in clinical practice. We aimed to investigate the prognostic value and associated clinical characteristics of subclinical pulmonary edema after acute STEMI without HF detected by chest computed tomography (CT).

**METHODS:** A total of 276 patients with acute STEMI without HF who underwent chest CT were included in this study. K-means clustering analysis was performed to classify the patients into different subgroups based on the mean lung density. Clinical characteristics of the different subgroups were compared and used to establish a machine learning model for discriminating between them. Relative risk (RR) for major adverse cardiovascular events (MACEs) during hospitalization was compared between the subgroups.

**RESULTS:** The patients were classified into two subgroups. Subgroup 2 showed higher mean lung density than subgroup 1 (median [IQR], −727 [−747, −704] vs. −806 [−826, −785] HU, P < 0.001), with significantly higher levels of cardiac enzymes and numbers of inflammatory cells and significantly worse left ventricular function than subgroup 1. In the model analysis, the most important clinical characteristics were the levels of cardiac enzymes, numbers of inflammatory cells, and left ventricular function. The risk for MACEs was higher in subgroup 2 than in subgroup 1 (RR, 2.12; P = 0.002).

**CONCLUSIONS:** Subclinical pulmonary edema after acute STEMI without HF was mainly associated with elevated levels of cardiac enzymes, followed by increased numbers of inflammatory cells and worse left ventricular function. In addition, subclinical pulmonary edema provided crucial prognostic information for patients during hospitalization.

## INTRODUCTION

Pulmonary edema is a severe complication in patients with acute myocardial infarction (MI) which indicates the development of heart failure (HF) and poor prognosis.^1,2^ Pulmonary edema has been shown to play important roles in risk stratification and short- and long-term prognoses in patients with acute MI.^1,3–5^ In acute MI without HF, there is a subclinical increase in pulmonary fluid content, possibly due to increased hydrostatic pressure and/or pulmonary capillary permeability.^6–8^ However, the prognostic value of subclinical pulmonary edema after acute MI without HF remains unclear.

Although chest X-ray and lung ultrasound are useful for the rapid qualitative or semiquantitative assessment of pulmonary edema in patients with acute MI,^4,5^ it is difficult to effectively assess subclinical pulmonary edema in those without HF. Chest computed tomography (CT) is a rapid and widely used tool that can sensitively detect minor alterations in lung density. Several previous studies have demonstrated that quantitative chest CT can effectively determine the severity of pulmonary edema and provide useful information for clinical evaluation in patients with HF.^9–11^ Recently, Jain et al. detected subclinical lung congestion using quantitative CT, providing new insights for the evaluation of pulmonary structure and function in patients with HF and preserved ejection fraction.^12^ To the best of our knowledge, no studies have evaluated subclinical pulmonary edema after acute MI without HF.

The mechanism of pulmonary edema after acute MI remains controversial. Most previous studies have revealed that pulmonary edema is caused by increased hydrostatic pressure due to left ventricular insufficiency and/or mitral regurgitation after acute MI.^13–15^ However, other animal and human studies have suggested that pulmonary inflammation and neural/hormonal mechanisms promote the development of pulmonary edema after acute MI.^6,16,17^

Therefore, we aimed to evaluate the prognostic value of subclinical pulmonary edema after acute MI without HF using quantitative chest CT. Moreover, we explored the clinical characteristics of patients associated with subclinical pulmonary edema.

## METHODS

### Study Population

This study was conducted at the Chest Hospital, Tianjin University in Tianjin, China. Consecutive patients with acute STEMI without HF who underwent chest CT between February 2015 and June 2021 were retrospectively enrolled. The inclusion criteria were patients who underwent chest CT and cardiac ultrasonography on admission. The exclusion criteria were as follows: a history of previous MI or coronary revascularization; history of pulmonary surgery; and patients with diseases that significantly alter lung density, such as lung tumors, emphysema, pulmonary tuberculosis, and interstitial fibrosis. This study was approved by the ethics committee of Chest Hospital, Tianjin University, and the requirement for written informed consent was waived for this retrospective study.

### Chest CT Acquisition Protocol

Chest CT scans (Emotion 16-slice CT scanner, Siemens Healthcare, Erlangen, Germany) were acquired on admission during full inspiration from the apex to lung base for all patients. The scanning parameters were as follows: tube voltage, 120 kV; smart mA tube current modulation; detector width, 1.5 mm; pitch, 1; matrix, 512 × 512; reconstruction slice thickness, 1.5 mm; and convolution kernel, sharp kernel (B70s).

### Clinical Characteristics

The clinical characteristics of patients, including hypertension, smoking, diabetes, dyslipidemia, heart rate, respiratory rate, blood routine, blood biochemistry, and cardiac ultrasonography results, were obtained by reviewing electronic medical records.

### Determination of the Mean Lung Density

The chest CT images were transferred to IntelliSpace Portal workstation (version 9.0, Philips Healthcare, Best, Netherlands) for postprocessing using chronic obstructive pulmonary disease analysis tool. During this process, the lungs were automatically segmented and the mean lung density was calculated. Manual correction was performed in case of inaccurate segmentation.

### Patient Subgroups

The sum of squared errors (SSE) and silhouette coefficient (SC) were used to determine the optimal number of subgroups based on the mean lung density in patients. The elbow point in the SSE curve and maximum value in the SC curve indicated the optimal cluster numbers.^18^ These two methods were performed using Python (Version 3.9.7). Subsequently, all patients were clustered into subgroups based on the corresponding cluster number using K-means clustering analysis algorithm. It is a widely used data-driven unsupervised machine learning algorithm owing to its simple principle and efficient operation.^19^ K-means clustering analysis was performed using IBM SPSS Statistics 26 (IBM Corporation, Armonk, NY, USA).

### Machine Learning

All clinical characteristics (except mean lung density) were used to establish a machine learning model (XGBoost) for discriminating patient subgroups. XGBoost is a novel boosted ensemble algorithm in the field of medicine, which exhibits high and reliable performance.^20^ This model assembles multiple weak classifiers and produces a robust classifier for improving prediction ability. First, all patients were randomly split into training (70%) and validation (30%) sets. Then, 10-fold cross-validation was used to optimize model parameters in the training set.^21^ Further, receiver operating characteristic (ROC) curves were used to evaluate the discriminative performance of the model based on the areas under the curves (AUCs). Finally, the ranking of clinical characteristics, according to their importance, for discriminating patient subgroups in the model was calculated.

### In-hospital Follow-up

The primary in-hospital outcome of this study was the occurrence of major adverse cardiovascular events (MACEs). MACEs include all-cause death, acute coronary syndrome (ACS), malignant arrhythmia (ventricular tachycardia, ventricular fibrillation, and III atrioventricular block), acute HF, cardiogenic shock, or stroke.

### Statistical Analysis

Statistical analysis was performed using IBM SPSS Statistics 26. One-sample Kolmogorov–Smirnov test was used to confirm a normal distribution. Normally distributed continuous variables were expressed as means ± standard deviations, whereas other variables were expressed as medians and interquartile ranges (IQRs). Categorical variables were expressed as counts (percentage). Student *t* test and Mann–Whitney *U* test were used to compare continuous variables with normal and non-normal distribution, respectively. Chi-square test was used to compare categorical variables. Spearman correlation analysis was used to evaluate the correlation between the mean lung density and clinical characteristics in both subgroups. Relative risk (RR) for MACE was compared between the subgroups. A two-sided *P* value of < 0.05 was considered to indicate statistical significance.

## RESULTS

### Patient Characteristics

Among 573 screened patients, 297 patients were excluded, as shown in Figure 1. Accordingly, the final population comprised 276 patients (221 men and 55 women; median age, 60 [IQR, 52, 67] years). Patient clinical characteristics are presented in Table 1.

**Figure 1.**
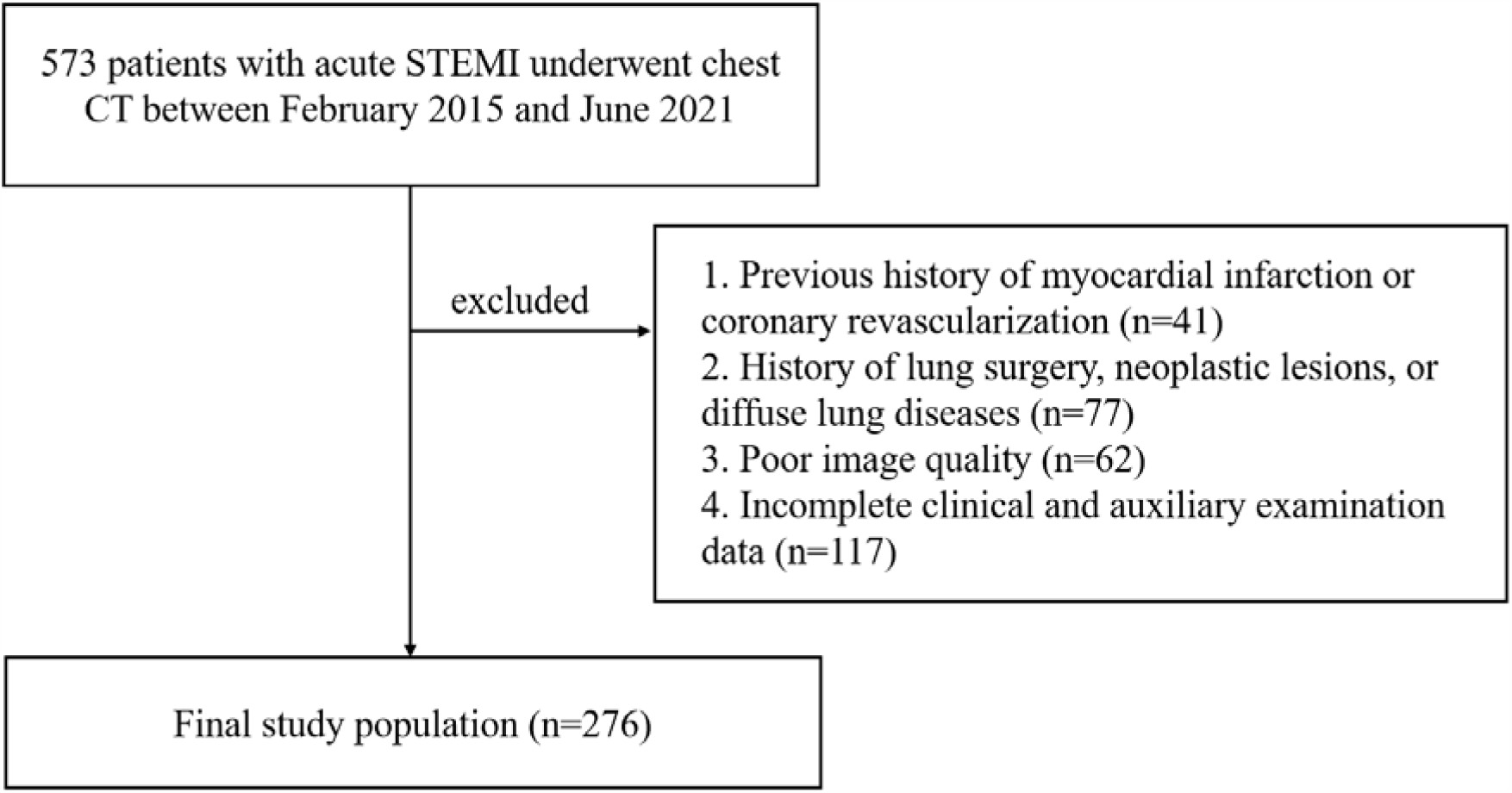
Flowchart of inclusion and exclusion criteria of the study. STEMI = ST-segment elevation myocardial infarction; CT = computed tomography.

**Table 1.**
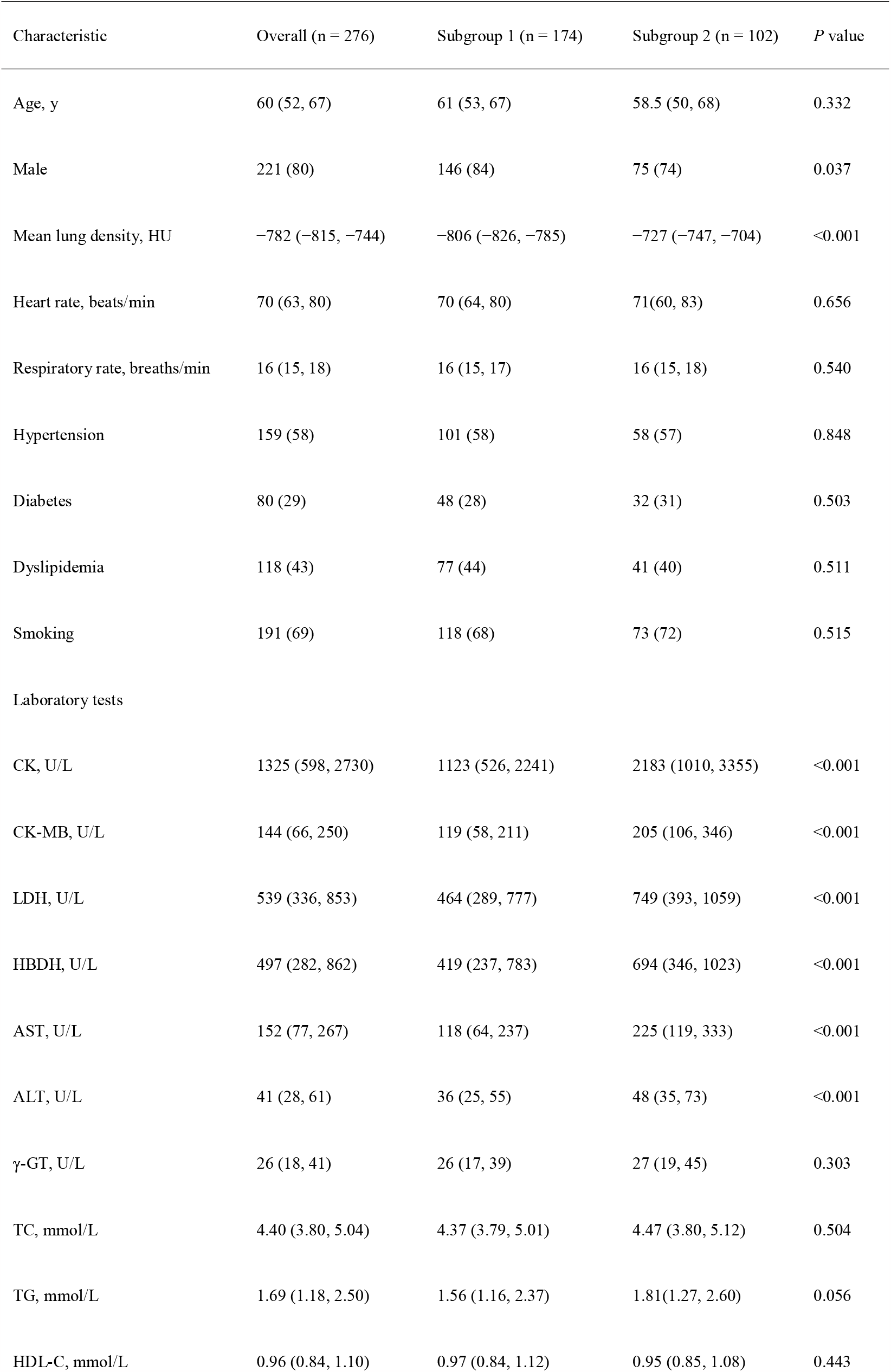
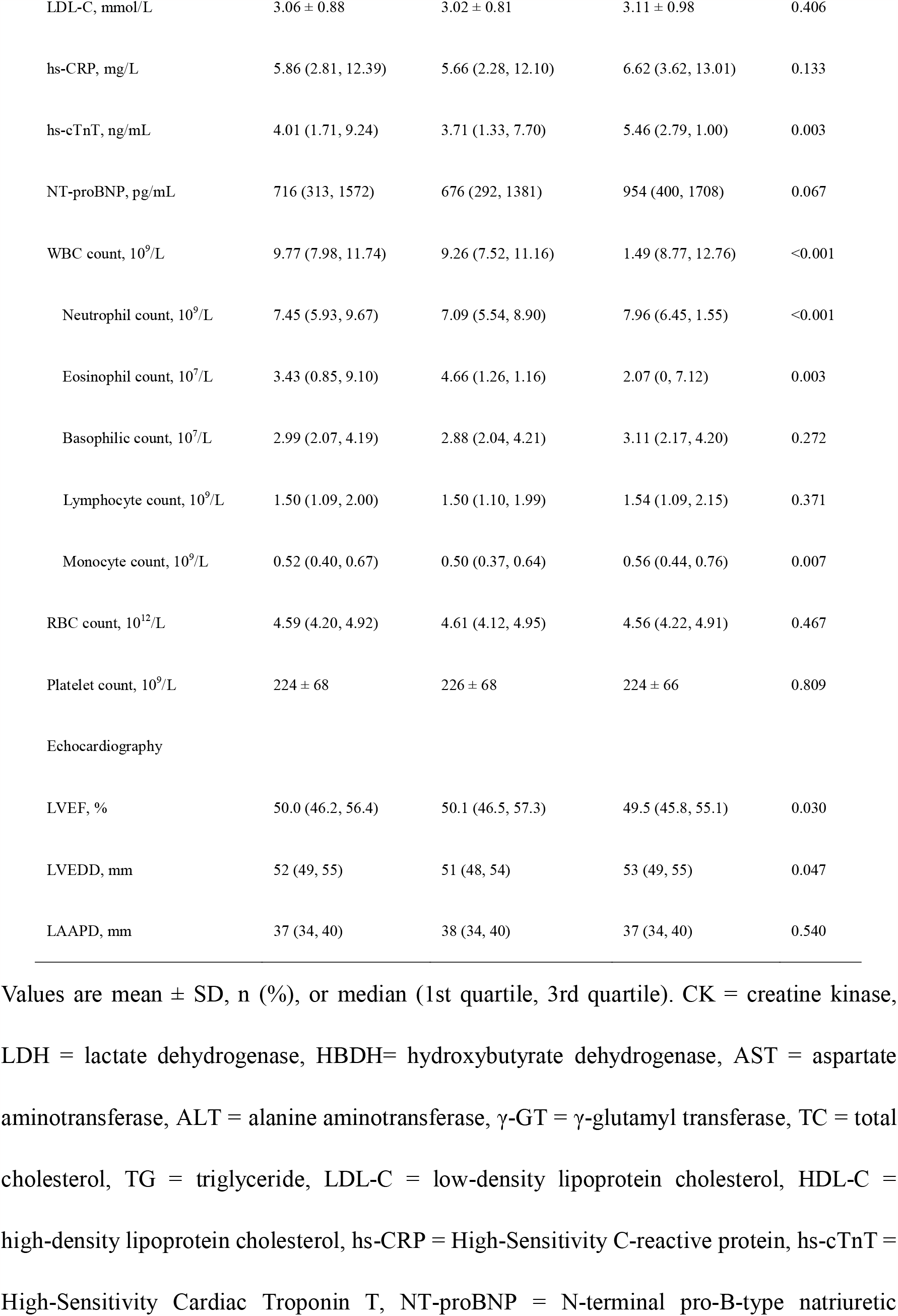
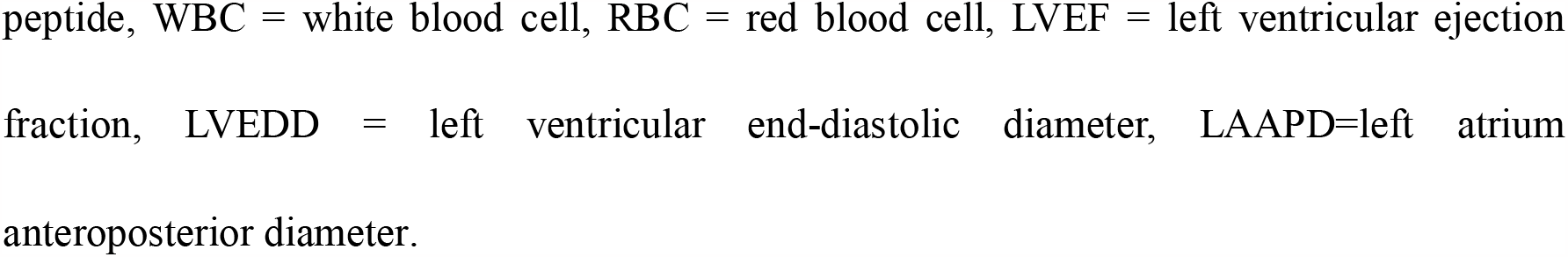
Clinical Characteristics of the Study Population.

| Characteristic | Overall (n = 276) | Subgroup 1 (n = 174) | Subgroup 2 (n = 102) | P value |
| --- | --- | --- | --- | --- |
| Age, y | 60 (52, 67) | 61 (53, 67) | 58.5 (50, 68) | 0.332 |
| Male | 221 (80) | 146 (84) | 75 (74) | 0.037 |
| Mean lung density, HU | -782 (-815, -744) | -806 (-826, -785) | -727 (-747, -704) | <0.001 |
| Heart rate, beats/min | 70 (63, 80) | 70 (64, 80) | 71(60, 83) | 0.656 |
| Respiratory rate, breaths/min | 16 (15, 18) | 16 (15, 17) | 16 (15, 18) | 0.540 |
| Hypertension | 159 (58) | 101 (58) | 58 (57) | 0.848 |
| Diabetes | 80 (29) | 48 (28) | 32 (31) | 0.503 |
| Dyslipidemia | 118 (43) | 77 (44) | 41 (40) | 0.511 |
| Smoking | 191 (69) | 118 (68) | 73 (72) | 0.515 |
| Laboratory tests |  |  |  |  |
| CK, U/L | 1325 (598, 2730) | 1123 (526, 2241) | 2183 (1010, 3355) | <0.001 |
| CK-MB, U/L | 144 (66, 250) | 119 (58, 211) | 205 (106, 346) | <0.001 |
| LDH, U/L | 539 (336, 853) | 464 (289, 777) | 749 (393, 1059) | <0.001 |
| HBDH, U/L | 497 (282, 862) | 419 (237, 783) | 694 (346, 1023) | <0.001 |
| AST, U/L | 152 (77, 267) | 118 (64, 237) | 225 (119, 333) | <0.001 |
| ALT, U/L | 41 (28, 61) | 36 (25, 55) | 48 (35, 73) | <0.001 |
| γ-GT, U/L | 26 (18, 41) | 26 (17, 39) | 27 (19, 45) | 0.303 |
| TC, mmol/L | 4.40 (3.80, 5.04) | 4.37 (3.79, 5.01) | 4.47 (3.80, 5.12) | 0.504 |
| TG, mmol/L | 1.69 (1.18, 2.50) | 1.56 (1.16, 2.37) | 1.81(1.27, 2.60) | 0.056 |
| HDL-C, mmol/L | 0.96 (0.84, 1.10) | 0.97 (0.84, 1.12) | 0.95 (0.85, 1.08) | 0.443 |
| LDL-C, mmol/L | 3.06 ± 0.88 | 3.02 ± 0.81 | 3.11 ± 0.98 | 0.406 |
| hs-CRP, mg/L | 5.86 (2.81, 12.39) | 5.66 (2.28, 12.10) | 6.62 (3.62, 13.01) | 0.133 |
| hs-cTnT, ng/mL | 4.01 (1.71, 9.24) | 3.71 (1.33, 7.70) | 5.46 (2.79, 1.00) | 0.003 |
| NT-proBNP, pg/mL | 716 (313, 1572) | 676 (292, 1381) | 954 (400, 1708) | 0.067 |
| WBC count, 10<sup>9</sup>/L | 9.77 (7.98, 11.74) | 9.26 (7.52, 11.16) | 1.49 (8.77, 12.76) | <0.001 |
| Neutrophil count, 10<sup>9</sup>/L | 7.45 (5.93, 9.67) | 7.09 (5.54, 8.90) | 7.96 (6.45, 1.55) | <0.001 |
| Eosinophil count, 10<sup>7</sup>/L | 3.43 (0.85, 9.10) | 4.66 (1.26, 1.16) | 2.07 (0, 7.12) | 0.003 |
| Basophilic count, 10<sup>7</sup>/L | 2.99 (2.07, 4.19) | 2.88 (2.04, 4.21) | 3.11 (2.17, 4.20) | 0.272 |
| Lymphocyte count, 10<sup>9</sup>/L | 1.50 (1.09, 2.00) | 1.50 (1.10, 1.99) | 1.54 (1.09, 2.15) | 0.371 |
| Monocyte count, 10<sup>9</sup>/L | 0.52 (0.40, 0.67) | 0.50 (0.37, 0.64) | 0.56 (0.44, 0.76) | 0.007 |
| RBC count, 10<sup>12</sup>/L | 4.59 (4.20, 4.92) | 4.61 (4.12, 4.95) | 4.56 (4.22, 4.91) | 0.467 |
| Platelet count, 10<sup>9</sup>/L | 224 ± 68 | 226 ± 68 | 224 ± 66 | 0.809 |
| Echocardiography |  |  |  |  |
| LVEF, % | 50.0 (46.2, 56.4) | 50.1 (46.5, 57.3) | 49.5 (45.8, 55.1) | 0.030 |
| LVEDD, mm | 52 (49, 55) | 51 (48, 54) | 53 (49, 55) | 0.047 |
| LAAPD, mm | 37 (34, 40) | 38 (34, 40) | 37 (34, 40) | 0.540 |

Values are mean ± SD, n (%), or median (1st quartile, 3rd quartile). CK = creatine kinase, LDH = lactate dehydrogenase, HBDH= hydroxybutyrate dehydrogenase, AST = aspartate aminotransferase, ALT = alanine aminotransferase, γ-GT = γ-glutamyl transferase, TC = total cholesterol, TG = triglyceride, LDL-C = low-density lipoprotein cholesterol, HDL-C = high-density lipoprotein cholesterol, hs-CRP = High-Sensitivity C-reactive protein, hs-cTnT = High-Sensitivity Cardiac Troponin T, NT-proBNP = N-terminal pro-B-type natriuretic
peptide, WBC = white blood cell, RBC = red blood cell, LVEF = left ventricular ejection fraction, LVEDD = left ventricular end-diastolic diameter, LAAPD=left atrium anteroposterior diameter.

### Subgrouping and Intersubgroup Differences in Clinical Characteristics

As shown in Figure 2A and B, the optimal cluster number was determined as two based on both SSE and SC curves. Overall, 176 (63%) and 102 (37%) patients were assigned to subgroup 1 and 2, respectively, using K-means clustering analysis based on their mean lung density. There were more men in subgroup 1 than in subgroup 2 (84% vs. 74%, *P* = 0.037). Subgroup 2 had higher mean lung density than subgroup 1 (median [IQR], −727 [−747, −704] vs. −806 [−826, −785] HU, *P* < 0.001). The distribution of both subgroups is presented in Figure 2C.

**Figure 2.**
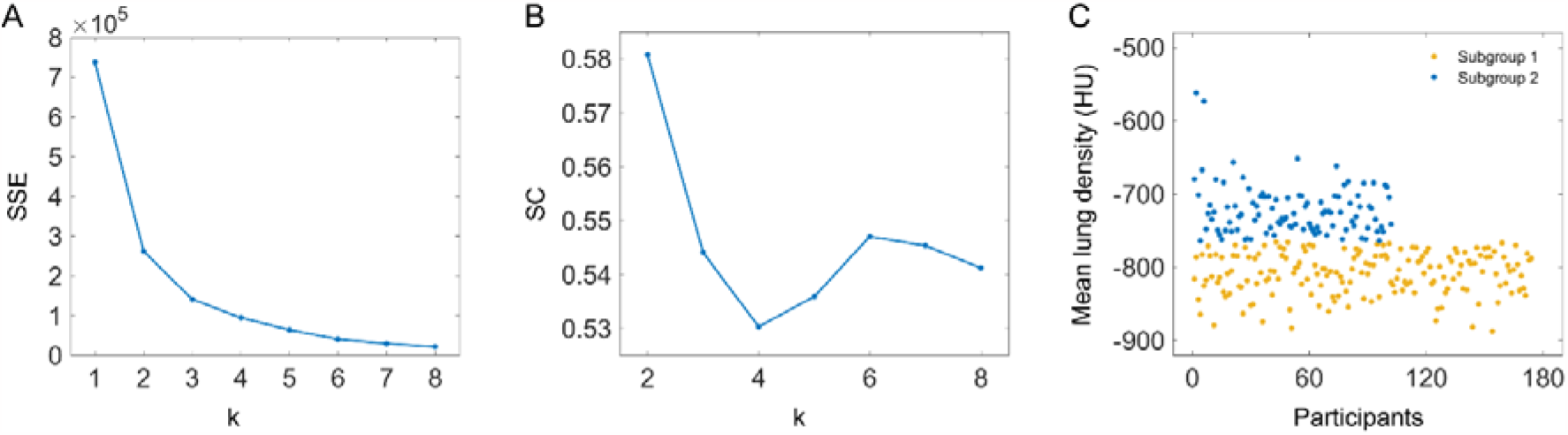
The sum of squared errors (SSE) and silhouette coefficient (SC) were used to determine the optimal number of subgroups based on the mean lung density (A, B). The optimal number of patient subgroups was determined as two based on these two methods. The two patient subgroups were classified using K-means clustering analysis based on the mean lung density (C). k = number of subgroups.

There were no significant differences in clinical characteristics, such as hypertension, diabetes, dyslipidemia, and smoking, between the subgroups. Based on laboratory tests, subgroup 2 showed significantly higher levels of creatine kinase (CK), CK-MB, lactate dehydrogenase (LDH), hydroxybutyrate dehydrogenase (HBDH), aspartate aminotransferase (AST), alanine aminotransferase (ALT), high-sensitivity cardiac troponin T (hs-cTnT) as well as higher counts of white blood cells (WBCs), neutrophils, and monocytes and lower counts of eosinophils than subgroup 1. Ultrasound cardiogram revealed that subgroup 2 had significantly lower left ventricular ejection fraction (LVEF) and higher left ventricular end-diastolic diameter than subgroup 1. Detailed clinical characteristics of the two subgroups are summarized in Table 1.

In correlation analysis, subgroup 1 exhibited a weak negative correlation between eosinophil and mean lung density; subgroup 2 exhibited relatively strong positive correlations among CK, CK-MB, AST, ALT, hs-cTnT, and mean lung density and negative correlations among monocytes, LVEF, and mean lung density. Compared with subgroup 1, the correlation between CK and mean lung density in subgroup 2 was significantly increased (r, 0.36 vs. 0.07; *P* = 0.014) (Table 2).

**Table 2.** Correlations between Clinical Characteristics and Mean Lung Density.

| Characteristic | Subgroup 1 (n = 174) |  | Subgroup 2 (n = 102) |  | *P* value |
| --- | --- | --- | --- | --- | --- |
|  | Correlation |  | Correlation |  |  |
|  | coefficient | *P* value | coefficient | P value |  |
| CK | 0.07 | 0.357 | 0.36 | <0.001 | 0.014 |
| CK-MB | 0.13 | 0.076 | 0.34 | <0.001 | 0.085 |
| LDH | 0.02 | 0.838 | 0.12 | 0.230 | 0.407 |
| HBDH | 0.02 | 0.822 | 0.13 | 0.177 | 0.348 |
| AST | 0.05 | 0.501 | 0.27 | 0.006 | 0.076 |
| ALT | 0.08 | 0.281 | 0.31 | 0.002 | 0.062 |
| hs-cTnT | 0.03 | 0.672 | 0.24 | 0.015 | 0.090 |
| WBC | 0.10 | 0.190 | 0.11 | 0.264 | 0.925 |
| Neutrophil | 0.09 | 0.238 | 0.15 | 0.145 | 0.655 |
| Eosinophil | − 0.17 | 0.023 | − 0.07 | 0.470 | 0.419 |
| Monocyte | − 0.10 | 0.191 | − 0.23 | 0.021 | 0.291 |
| LVEF | − 0.07 | 0.352 | − 0.20 | 0.044 | 0.299 |
| LVEDD | 0.13 | 0.085 | 0.03 | 0.765 | 0.421 |

CK = creatine kinase, LDH = lactate dehydrogenase, HBDH= hydroxybutyrate dehydrogenase, AST = aspartate aminotransferase, ALT = alanine aminotransferase, hs-cTnT = High-Sensitivity Cardiac Troponin T, WBC = white blood cell, RBC = red blood cell, LVEF = left ventricular ejection fraction, LVEDD = left ventricular end-diastolic diameter.

### Machine Learning for Discriminating the Subgroups

The XGBoost model was established for discriminating the subgroups based on 33 clinical characteristics. In this machine learning model, the AUCs of the training and test sets were 0.81 and 0.73, respectively (Figure 3). The ranking of clinical characteristics, according to their importance, for discriminating subgroups in the XGBoost model is shown in Figure 4. Of the clinical characteristics, CK level was the most important, followed by AST and LDH levels. Similarly, neutrophils, LVEF, WBC, and CK-MB showed relatively high importance. However, the high ranking of total cholesterol (TC) and low-density lipoprotein cholesterol (LDL-C) was unexpected.

**Figure 3.**
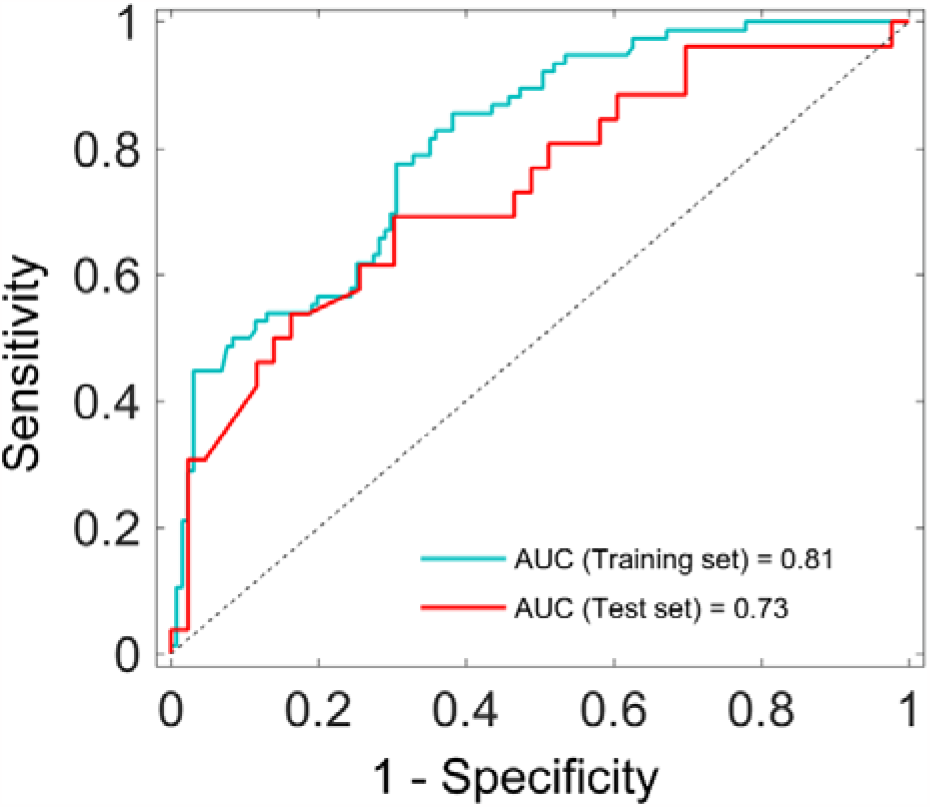
Performance of the XGBoost model in discriminating subgroups in the training and test sets using 33 clinical characteristics. AUC = area under the curve.

**Figure 4.**
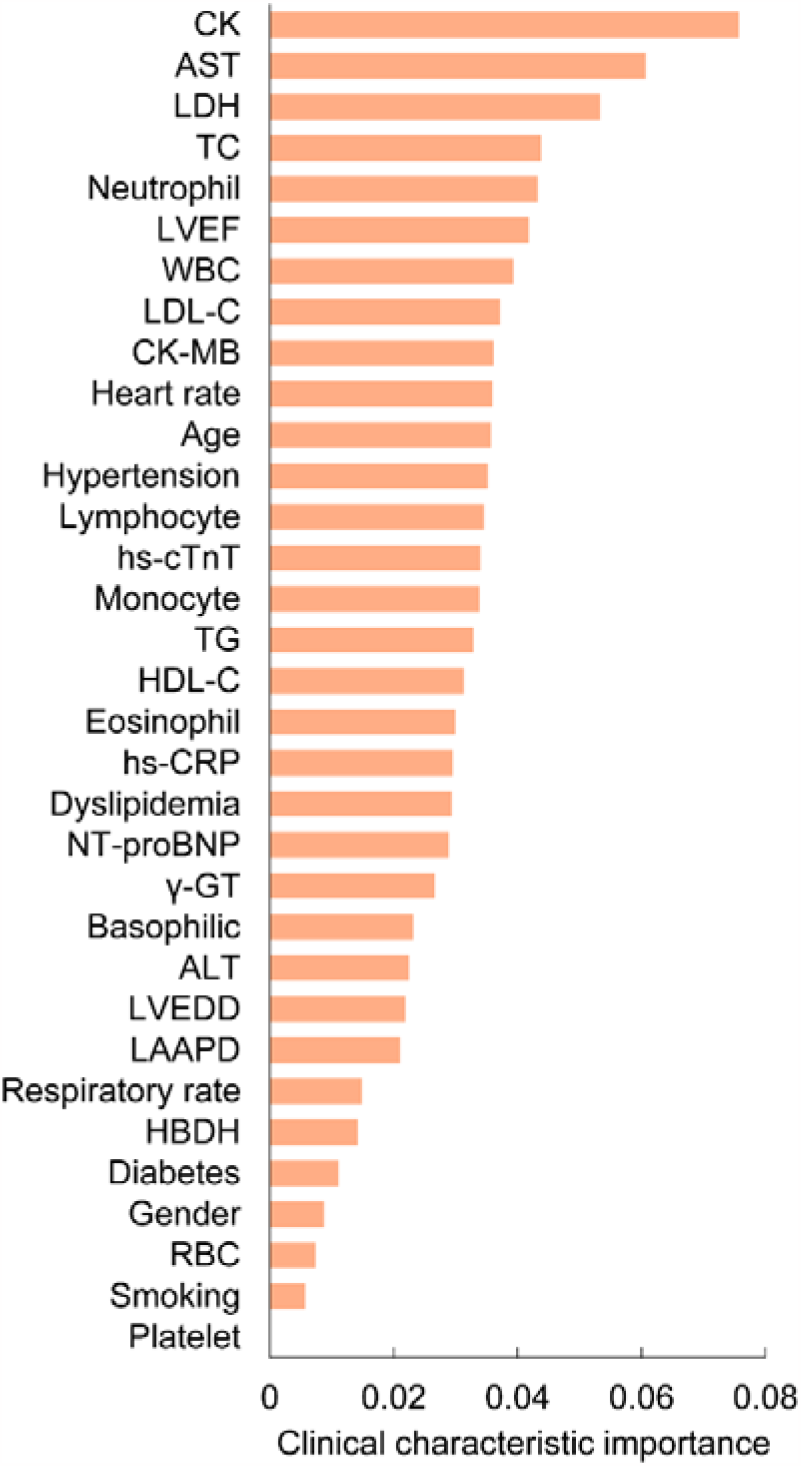
The ranking of 33 clinical characteristics, according to their importance, in the XGBoost model. CK = creatine kinase, LDH = lactate dehydrogenase, HBDH= hydroxybutyrate dehydrogenase, AST = aspartate aminotransferase, ALT = alanine aminotransferase, γ-GT = γ-glutamyl transferase, TC = total cholesterol, TG = triglyceride, LDL-C = low-density lipoprotein cholesterol, HDL-C = high-density lipoprotein cholesterol, hs-CRP = High-Sensitivity C-reactive protein, hs-cTnT = High-Sensitivity Cardiac Troponin T, NT-proBNP = N-terminal pro-B-type natriuretic peptide, WBC = white blood cell, RBC = red blood cell, LVEDD = left ventricular end-diastolic diameter, LVEF = left ventricular ejection fraction.

### In-hospital Outcome

In total, 56 patients (2.3%) suffered from MACEs during hospitalization, including 24 acute HF cases, 17 malignant arrhythmias cases, 10 ACS cases, 4 cardiogenic shock cases, and 1 death. MACEs occurred in 25 patients (14.4%) in subgroup 1 and 31 patients (3.4%) in subgroup 2. The risk for MACE was significantly higher in subgroup 2 than in subgroup 1 during hospitalization (RR, 2.12; *P* = 0.002).

## DISCUSSION

In this study, machine learning based on quantitative chest CT data was used to stratify the risk of patients with acute STEMI without HF and explore their associated clinical characteristics. Patients were divided into two subgroups: subgroup 1 comprising patients with mild subclinical pulmonary edema and subgroup 2 consisting of patients with relatively severe subclinical pulmonary edema. We revealed that subclinical pulmonary edema was mainly associated with elevated levels of cardiac enzymes (especially CK), followed by increased numbers of inflammatory cells and worse left ventricular function. Further, subgroup 2 had a significantly higher risk for MACEs during hospitalization than subgroup 1 (RR, 2.12; *P* = 0.002).

From a clinical perspective, our results showed that subclinical pulmonary edema in patients with acute STEMI without HF was mainly associated with high levels of cardiac enzymes, particularly CK. The elevated levels of cardiac enzymes indicate myocardial injury; this was commonly used for the diagnosis of acute MI.^22,23^ Currently, although these cardiac enzymes have been replaced by the more sensitive and specific indicator, i.e., cTnT,^24^ CK and CK-MB still play important roles in assessing the infarct size and prognosis of patients with MI after primary percutaneous coronary intervention;^25,26^ moreover, they are the therapeutic targets of myocardial ischemia–reperfusion injury.^27^ In this study, the close association between cardiac enzymes and subclinical pulmonary edema revealed the potential clinical value of cardiac enzymes in patients with STEMI. To the best of our knowledge, no relevant data is available and further research is warranted to explore the specific mechanism underlying this association. In addition, we revealed that inflammatory cells, particularly neutrophils, after acute STEMI may play an important role in the formation of subclinical pulmonary edema. This finding has been supported by several previous studies.^6,16,17^ In particular, Wu et al. noted marked pulmonary edema in rats with acute MI, with significantly increased pulmonary neutrophil infiltration. A possible mechanism is that inflammation damages the endothelial cells of pulmonary capillaries, resulting in increased permeability. Unexpectedly, the contribution of left ventricular dysfunction in the formation of subclinical pulmonary edema was relatively weak in our study. Although this finding contradicts that of previous studies showing that pulmonary edema is mainly caused by left ventricular dysfunction,^14,28^ our results may explain an unanswered question as to why pulmonary edema frequently occurs in cases of mild left ventricular dysfunction.^29^

Early risk stratification in patients with acute STEMI is critical for their management during hospitalization. Our study revealed that patients with relatively severe subclinical pulmonary edema had a significantly higher risk (RR, 2.12; *P* = 0.002) for MACEs during hospitalization than those with mild subclinical pulmonary edema. Previous studies have indicated the clinical importance of pulmonary edema in acute MI. For instance, initial chest X-ray can provide information regarding risk stratification and prognosis,^30^ and admission lung ultrasound along with Killip classification was more sensitive than physical examination to identify patients with a high risk for in-hospital mortality.^31^ Due to the limitations of these techniques, the prognostic value of subclinical pulmonary edema after acute STEMI without HF remains unexplored. Our study addressed this limitation using quantitative chest CT and highlighted the prognostic value of subclinical pulmonary edema for risk stratification in patients with acute STEMI without HF.

### Limitations

There are several limitations in our study. First, this was a retrospective study and data were collected on admission, either due to atypical symptoms or to exclude other possible diseases. Chest CT is not a routine examination in patients with acute MI, which may delay its diagnosis and treatment, thereby limiting further research and clinical practice. Second, our study only found close associations between subclinical pulmonary edema and clinical characteristics, but the specific pathophysiological mechanisms underlying these associations remain unclear. Third, the long-term prognostic value of subclinical pulmonary edema was not investigated in this study as the relevant data were not available, thus warranting further research.

## Conclusions

Chest CT was used to detect and assess subclinical pulmonary edema after STEMI without HF in this study. Subclinical pulmonary edema was mainly associated with elevated levels of cardiac enzymes, followed by increased numbers of inflammatory cells and worse left ventricular function; however, the specific pathophysiological mechanisms of this condition require further exploration. Risk stratification based on subclinical pulmonary edema showed strong prognostic value for patients during hospitalization and may provide new insights into clinical practice.

## Data Availability

The data underlying this article will be shared on reasonable request to the corresponding author.

## Acknowledgments

This research was supported by the Tianjin Health Science and technology project (MS20015). The authors acknowledge the support of the Tianjin Key Laboratory of Cardiovascular Emergency and Critical Care, Tianjin Municipal Science and Technology Bureau.

## References

1. Killip T 3rd, Kimball JT. Treatment of myocardial infarction in a coronary care unit. A two year experience with 250 patients. Am J Cardiol. 1967;20(4):457–464. doi: 10.1016/0002-9149(67)90023-9

2. Velazquez EJ, Francis GS, Armstrong PW, et al. An international perspective on heart failure and left ventricular systolic dysfunction complicating myocardial infarction: the VALIANT registry. Eur Heart J. 2004;25(21):1911–1919. doi: 10.1016/j.ehj.2004.08.006

3. Warnowicz MA, Parker H, Cheitlin MD. Prognosis of patients with acute pulmonary edema and normal ejection fraction after acute myocardial infarction. Circulation. 1983;67(2):330–334. doi: 10.1161/01.cir.67.2.330

4. He J, Yi S, Zhou Y, et al. B-Lines by Lung Ultrasound Can Predict Worsening Heart Failure in Acute Myocardial Infarction During Hospitalization and Short-Term Follow-Up. Front Cardiovasc Med. 2022;9:895133. doi: 10.3389/fcvm.2022.895133

5. Gottlieb S, Moss AJ, McDermott M, Eberly S. Interrelation of left ventricular ejection fraction, pulmonary congestion and outcome in acute myocardial infarction. Am J Cardiol. 1992;69(12):977–984. doi: 10.1016/0002-9149(92)90850-x

6. Guazzi M, Arena R, Guazzi MD. Evolving changes in lung interstitial fluid content after acute myocardial infarction: mechanisms and pathophysiological correlates. Am J Physiol Heart Circ Physiol. 2008;294(3):H1357–H1364. doi: 10.1152/ajpheart.00866.2007

7. Slutsky RA, Peck WW, Higgins CB. Pulmonary edema formation with myocardial infarction and left atrial hypertension: intravascular and extravascular pulmonary fluid volumes. Circulation. 1983;68(1):164–169. doi: 10.1161/01.cir.68.1.164

8. Luz da PL, Shubin H, Weil MH, Jacobson E, Stein L. Pulmonary edema related to changes in colloid osmotic and pulmonary artery wedge pressure in patients after acute myocardial infarction. Circulation. 1975;51(2):350–357. doi: 10.1161/01.cir.51.2.350

9. Barile M, Hida T, Hammer M, Hatabu H. Simple quantitative chest CT for pulmonary edema. Eur J Radiol Open. 2020;7:100273. doi: 10.1016/j.ejro.2020.100273

10. Chase SC, Taylor BJ, Cross TJ, Coffman KE, Olson LJ, Johnson BD. Influence of Thoracic Fluid Compartments on Pulmonary Congestion in Chronic Heart Failure. J Card Fail. 2017;23(9):690–696. doi: 10.1016/j.cardfail.2017.07.394

11. Morooka N, Watanabe S, Masuda Y, Inagaki Y. Estimation of pulmonary water distribution and pulmonary congestion by computed tomography. Jpn Heart J. 1982;23(5):697–709. doi: 10.1536/ihj.23.697

12. Jain CC, Tschirren J, Reddy YNV, Melenovsky V, Redfield M, Borlaug BA. Subclinical Pulmonary Congestion and Abnormal Hemodynamics in Heart Failure With Preserved Ejection Fraction. JACC Cardiovasc Imaging. 2022;15(4):629–637. doi: 10.1016/j.jcmg.2021.09.017

13. Finsen AV, Christensen G, Sjaastad I. Echocardiographic parameters discriminating myocardial infarction with pulmonary congestion from myocardial infarction without congestion in the mouse. J Appl Physiol (1985). 2005;98(2):680–689. doi: 10.1152/japplphysiol.00924.2004

14. Gong FF, Vaitenas I, Malaisrie SC, Maganti K. Mechanical Complications of Acute Myocardial Infarction: A Review. JAMA Cardiol. 2021;6(3):341–349. doi: 10.1001/jamacardio.2020.3690

15. Bursi F, Enriquez-Sarano M, Nkomo VT, et al. Heart failure and death after myocardial infarction in the community: the emerging role of mitral regurgitation. Circulation. 2005;111(3):295–301. doi: 10.1161/01.CIR.0000151097.30779.04

16. Wu J, Guo Z, Wang LL, Zhang RL. Degeneration of sensory afferent nerves enhances pulmonary inflammatory alterations in acute myocardial infarction in rats. Cardiovasc Pathol. 2012;21(3):149–157. doi: 10.1016/j.carpath.2011.05.001

17. Ben Driss A, Devaux C, Henrion D, et al. Hemodynamic stresses induce endothelial dysfunction and remodeling of pulmonary artery in experimental compensated heart failure. Circulation. 2000;101(23):2764–2770. doi: 10.1161/01.cir.101.23.2764

18. Kodinariya TM, Makwana PR. Review on determining number of Cluster in K-Means Clustering. International Journal. 2013;1:90–95.

19. Jain AK, Dubes RC. Algorithms for clustering data. Prentice-Hall, Inc.. 1988.

20. Chen T, Guestrin C. Xgboost: A scalable tree boosting system; proceedings of the Proceedings of the 22nd acm sigkdd international conference on knowledge discovery and data mining. New York, NY, USA. 2016:785.

21. Kim J-H. Estimating classification error rate: Repeated cross-validation, repeated hold-out and bootstrap. Comput Stat Data Anal. 2009;53:3735–3745. doi: 10.1016/j.csda.2009.04.009

22. Danese E, Montagnana M. An historical approach to the diagnostic biomarkers of acute coronary syndrome. Ann Transl Med. 2016;4(10):194. doi: 10.21037/atm.2016.05.19

23. Smith AF, Radford D, Wong CP, Oliver MF. Creatine kinase MB isoenzyme studies in diagnosis of myocardial infarction. Br Heart J. 1976;38(3):225–232. doi: 10.1136/hrt.38.3.225

24. Alpert JS, Thygesen K, Antman E, Bassand JP. Myocardial infarction redefined--a consensus document of The Joint European Society of Cardiology/American College of Cardiology Committee for the redefinition of myocardial infarction. J Am Coll Cardiol. 2000;36(3):959–969. doi: 10.1016/s0735-1097(00)00804-4

25. Nienhuis MB, Ottervanger JP, de Boer MJ, et al. Prognostic importance of creatine kinase and creatine kinase-MB after primary percutaneous coronary intervention for ST-elevation myocardial infarction. Am Heart J. 2008;155(4):673–679. doi: 10.1016/j.ahj.2007.11.004

26. Turer AT, Mahaffey KW, Gallup D, et al. Enzyme estimates of infarct size correlate with functional and clinical outcomes in the setting of ST-segment elevation myocardial infarction. Curr Control Trials Cardiovasc Med. 2005;6(1):12. doi: 10.1186/1468-6708-6-12

27. Cao F, Zervou S, Lygate CA. The creatine kinase system as a therapeutic target for myocardial ischaemia-reperfusion injury. Biochem Soc Trans. 2018;46(5):1119–1127. doi: 10.1042/BST20170504

28. Spencer FA, Meyer TE, Gore JM, Goldberg RJ. Heterogeneity in the management and outcomes of patients with acute myocardial infarction complicated by heart failure: the National Registry of Myocardial Infarction. Circulation. 2002;105(22):2605–2610. doi: 10.1161/01.cir.0000017861.00991.2f

29. Møller JE, Pellikka PA, Hillis GS, Oh JK. Prognostic importance of diastolic function and filling pressure in patients with acute myocardial infarction. Circulation. 2006;114(5):438–444. doi: 10.1161/CIRCULATIONAHA.105.601005

30. Battler A, Karliner JS, Higgins CB, et al. The initial chest x-ray in acute myocardial infarction. Prediction of early and late mortality and survival. Circulation. 1980;61(5):1004–1009. doi: 10.1161/01.cir.61.5.1004

31. Araujo GN, Silveira AD, Scolari FL, et al. Admission Bedside Lung Ultrasound Reclassifies Mortality Prediction in Patients With ST-Segment-Elevation Myocardial Infarction. Circ Cardiovasc Imaging. 2020;13(6):e010269. doi: 10.1161/CIRCIMAGING.119.010269

